# Understand research hotspots surrounding COVID-19 and other coronavirus infections using topic modeling

**DOI:** 10.1101/2020.03.26.20044164

**Authors:** Mengying Dong, Xiaojun Cao, Mingbiao Liang, Lijuan Li, Guangjian Liu, Huiying Liang

## Abstract

**Background:** Severe acute respiratory syndrome coronavirus 2 (SARS-CoV-2) is a virus that causes severe respiratory illness in humans, which results in global outbreak of novel coronavirus disease (COVID-19) currently. This study aimed to evaluate the characteristics of publications involving coronaviruses as well as COVID-19 by using topic modeling.

**Methods:** We extracted all abstracts and retained the most informative words from the COVID-19 Open Research Dataset, which contains 35,092 pieces of coronavirus related literature published up to March 20, 2020. Using Latent Dirichlet Allocation modeling, we trained a topic model from the corpus, analyzed the semantic relationships between topics and compared the topic distribution between COVID-19 and other CoV infections.

**Results:** Eight topics emerged overall: clinical characterization, pathogenesis research, therapeutics research, epidemiological study, virus transmission, vaccines research, virus diagnostics, and viral genomics. It was observed that current COVID-19 research puts more emphasis on clinical characterization, epidemiological study, and virus transmission. In contrast, topics about diagnostics, therapeutics, vaccines, genomics and pathogenesis only account for less than 10% or even 4% of all the COVID-19 publications, much lower than those of other CoV infections.

**Conclusions:** These results identified knowledge gaps in the area of COVID-19 and offered directions for future research.

## Introduction

Coronaviruses (CoVs) are enveloped, positive single-stranded large RNA viruses that cause respiratory and intestinal infections in animals and humans (1). To date, there have seven human coronaviruses (HCoVs) been identified. The alpha-CoVs HCoVs-NL63 and HCoVs-229E and the beta-CoVs HCoVs-OC43, HCoVs-HKU1 only cause mild respiratory illness (2, 3). However, in the past two decades, two novel coronaviruses, severe acute respiratory syndrome CoV (SARS-CoV) and Middle East respiratory syndrome CoV (MERS-CoV), caused severe human diseases in local countries and regions, representing fatality rates of around 10% and 35%, respectively (4-6). Now, the seventh HCoV, SARS-CoV-2 (formerly 2019-nCoV), is causing an outbreak of coronavirus disease (COVID-19) all over the world.

Compared to the earlier epidemics by CoVs, COVID-19 is highly contagious and has been reported with more than 1,000,000 confirmed cases as on April 4, 2020 (7). Given the spread of the new CoV and its impacts on human health, governments are facing increasing pressure to stop the COVID-19 pandemic. The White House and a coalition of leading research groups have prepared a COVID-19 Open Research Dataset (CORD-19) and issued a call to action to the world’s artificial intelligence experts to support the ongoing fight against this infectious disease. The research community has also responded rapidly and substantial literature about this epidemic has emerged.

Understanding of COVID-19 is evolving rapidly. It is essential to understand the emerging scientific knowledge to coordinate COVID-19 research globally. Recently, the WHO has published a Global Research Roadmap with immediate, mid-term and longer-term priorities to enable the implementation of priority research (8). Bonilla-Aldana DK et al. (9) and Md Mahbub Hossain MBBS (10) have performed bibliometric analysis to evaluate the scientific literature on coronavirus infections as well as COVID-19, basing on indicators such as the number of articles, the productivity of authors, geographic distribution of articles and prominent keywords. However, to the best of our knowledge, there is still no quantitative thematic analysis available to date that focuses on CoVs.

Addressing this knowledge gap requires not only elaboration at the text level but also an understanding of semantic structures. Top modeling is the most notable approach for analyzing text to identify semantic content (11). Its objective is to find latent semantic topics within vast and unstructured collections of literature. Among several topic modeling algorithms, Latent Dirichlet Allocation (LDA) is the most fundamental one and has been shown to be effective at discovering semantic structures and distinct topics from a corpus (12-18). This kind of data-driven analysis can generally shift the information overload burden from researchers to the computer via algorithmic and statistical methods.

The purpose of this work was to conduct LDA modeling for semantic and quantitative evaluations of the current status of literature on CoV infections as well as COVID-19, identify broad research topics and how these topics interact with one another. More importantly, this work can benefit COVID-19 research coordination by recognizing high priority scientific topics. We found that topics of clinical characterization, epidemiology, and virus transmission are hotspots for COVID-19 at present, while research on pathogenesis, therapeutics, virus diagnostics, vaccines and viral genomics are urgently needed.

## Methods

### Corpus

Data used in this study were obtained from CORD-19, which was prepared by the White House and a coalition of leading research groups in response to the COVID-19 pandemic (19). CORD-19 contains all research about COVID-19, SARS-CoV-2, and other CoVs (e.g. SARS, MERS, etc.) up to March 20, 2020, including over 44,000 scholarly articles, from the following sources: 1) PubMed’s PMC open access corpus; 2) a corpus maintained by the WHO; and 3) bioRxiv and medRxiv pre-prints.

Abstracts were firstly exported from CORD-19 and merged into a local repository for further processing. Next, 8,420 records with an empty or non-English abstract (i.e., not provided in the database) were removed from the corpus. Then, 708 duplicates were removed based on the title (stripped of all special characters and spaces) or the digital object identifier (DOI, if available). The final corpus contained 35,092 abstracts of all coronaviruses-related articles. We further retrieved COVID-19 data by administering the query, “COVID” OR “COVID-19” OR “2019-nCoV” OR “SARS-CoV-2” OR “Novel coronavirus”, and limiting the publication time to 2020. In total, 1,482 articles were identified as COVID-19-related research.

This study is a retrospective analysis of accepted and publicly disclosed research, which does not meet the definition of human research under the US Department of Health and Human Services protection of human subject regulations. Data were not deidentified and are publicly available. There was no intervention, interaction with researchers, or use of private information to perform the analyses in this study.

### Data pre-processing

We applied a standard approach for tokenization, lemmatization and part-of-speech tagging to pre-process the biomedical texts. We also removed the following words to get the clean corpus: 1) punctuation; 2) digits; 3) special characters; 4) words that appeared fewer than 5 times in the corpus; 5) useless words from text collection, including coordinating conjunctions, cardinal numbers, prepositions, pronouns and so on; 6) common terms in scientific articles, e.g., ‘method’ and ‘introduction’, as well as ‘virus’, which is thought to appear in most abstracts.

### Topic modeling

We built a topic model using LDA (20, 21) from the article abstracts within the corpus. LDA assumes individual document as random mixtures over latent topics, with topics in turn being probability distributions over multiple vocabularies, and topics being uncorrelated. The Gibbs sampling algorithm is used to estimate the topic distribution parameters of the document and the multi-distribution of the vocabulary on each topic. The evaluation index in the statistical language model, coherence score, was used to find the optimal number of topics in the corpus. The coherence score is calculated by the co-occurrence frequency of the words in the sliding window, which increases with the increase of sentence similarity, meaning the higher, the better. Although the coherence score can help to decide the most appropriate topic number, such number sometimes can be still large for manually understanding. Under this circumstance, manual examination should be performed to evaluate models with different numbers of topics with the aid of expert knowledge to thoroughly understand the corpus. For each model, 15 top words per topic were examined to assess scientific coherence of the words as a set, overlap in topic words across topics, and human understandability. The selected model was used for all subsequent analyses. LDA modeling is performed and visualized on the entire corpus via the Python language.

## Results

### Word-level analysis

Before profiling topics in the corpus, a co-occurrence map of the 50 most frequent words in the abstracts were generated to describe the research hotspots of coronaviruses from a macro-level viewpoint (Fig. 1). The top ten co-occurring words were ‘infection’ (n = 49753), ‘cell’ (n = 36689), ‘protein’ (n = 26593), ‘diseases’ (n = 19643), ‘patient’ (n = 19307), ‘human’ (n = 13773), ‘respiratory’ (n = 12572), ‘response’ (n = 12473), ‘gene’ (n = 11138), and ‘identify’ (n = 11100). It is obvious that most of these words take ‘infection’ as the central node, depicting the virus attack event from clinical, macroscopic and microscopic viewpoints. Certainly, such map treats each word as a topic and gets the relationships between words simply basing on co-occurrence statistics. Its granularity is too small to capture the semantic information of CoV research.

**Fig 1.**
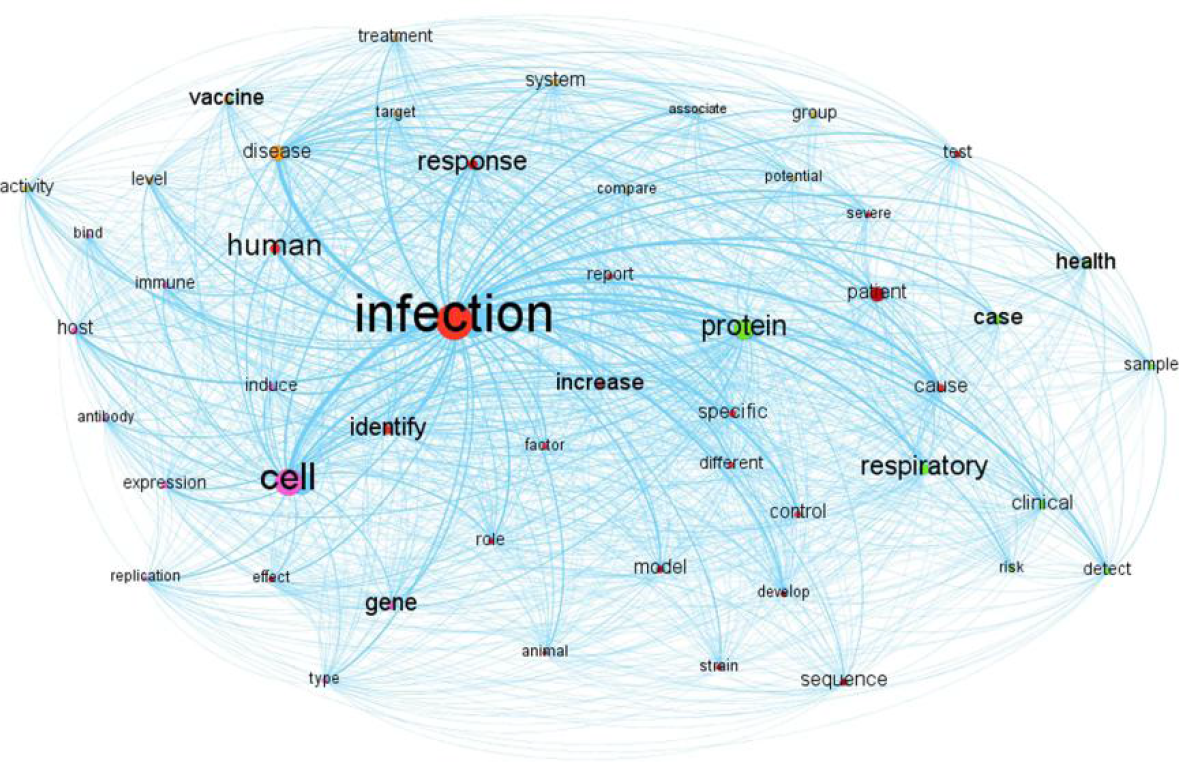
Co-occurrence map of the 50 most frequent words.

### Topic modeling analysis

We then employed LDA modeling to reveal the latent intellectual topics in the literature corpus based on the well-chosen words of the article abstracts (see Methods). After careful manual evaluation of the topics and their top words, the model fitting eight topics was selected based on topic coherence and human understanding. The final model is visualized as a bubble graph in Fig 2, with one bubble representing one topic and the size indicating its popularity. As shown in the figure, all the bubbles are completely separate from each other, suggesting the suitability of the model. Increasing the number of topics would make each individual topic more specific and might increase overlap between topics. Decreasing the number of topics would result in topics to be more high-level abstract.

**Fig 2.**
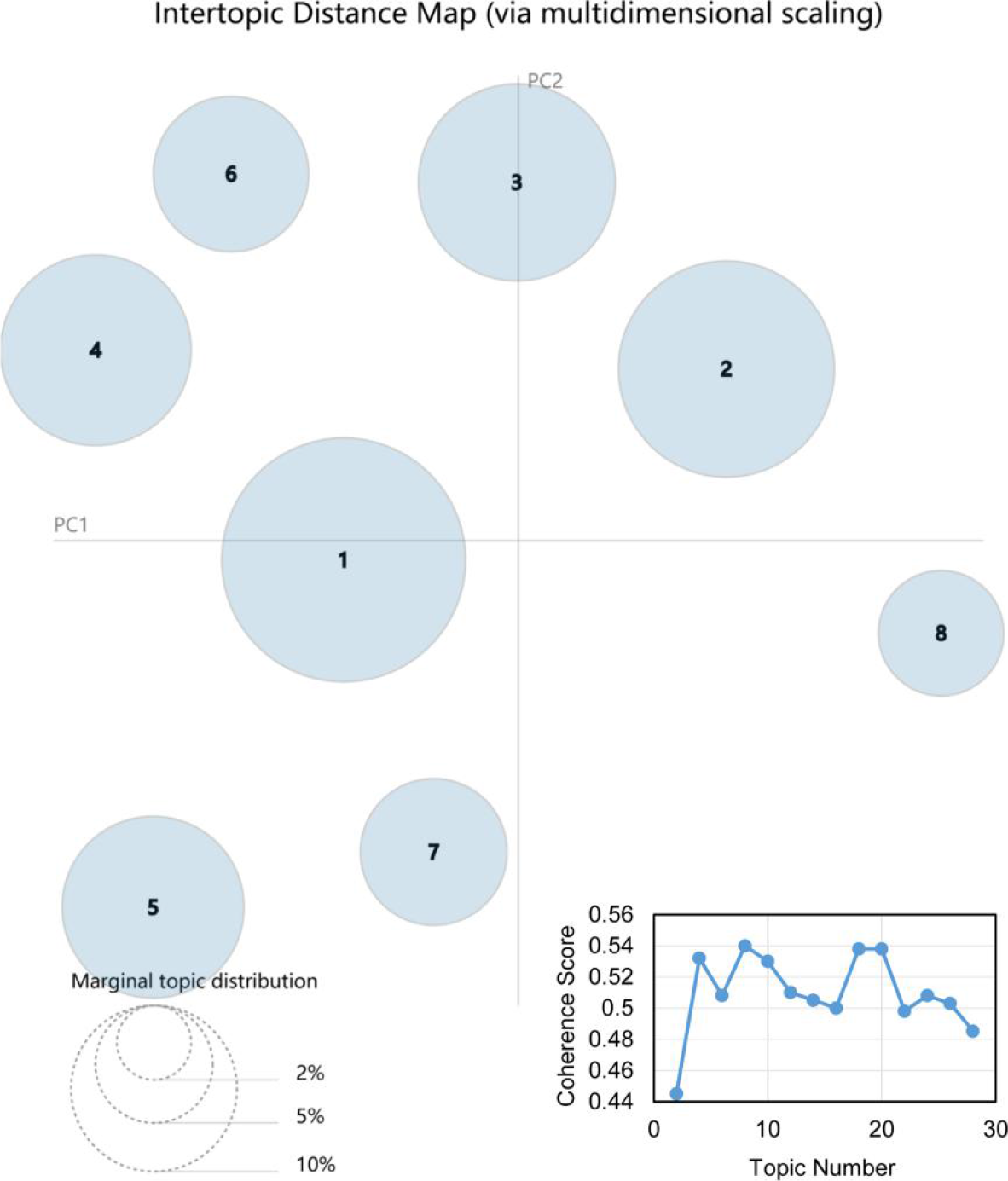
Visualization of the eight-topic model trained on the whole corpus. One bubble represents one topic. The topics are ordered based on prevalence in the corpus. Inset: the coherence scores of models with different numbers of topics.

We assigned a potential theme to each topic by manual examinations based on semantics analysis of representative words in each topic. Table 1 shows the 15 most frequent words for each of the eight topics. In most cases, topics were easily recognizable representing specific subjects about the viruses, or the disease, or the public health and so on. The first most dominant topic was enriched for the clinical characterization, with words such as ‘infection’, ‘cause’, ‘disease’, ‘severe’, ‘respiratory’, ‘acute’, ‘child’ and ‘symptom’. Representative words of topic 2 include ‘cell’, ‘protein’, ‘expression’, ‘bind’, ‘replication’, ‘activity’ and ‘membrane’, which usually are used to describe micromolecular reaction. We labeled this topic ‘pathogenesis research’. Topic 3 is focused on the therapeutics research against viruses, with words such as ‘target’, ‘novel’, ‘potential’, ‘antiviral’, ‘new’, ‘drug’, ‘therapeutic’ and ‘development’. This topic contains some noise, because the words ‘host’ and ‘specie’ might involve in the animal and environmental research rather than therapeutics. Topic 4 is concerned with the epidemiological study with related words such as ‘health’, ‘outbreak’, ‘model’, ‘epidemic’, ‘public’, ‘spread’, ‘global’, ‘emerge’ and ‘surveillance’. Topics 5 to 8 obviously address topics regarding virus transmission, vaccines research, virus diagnostics and viral genomics, respectively.

**Table 1.**
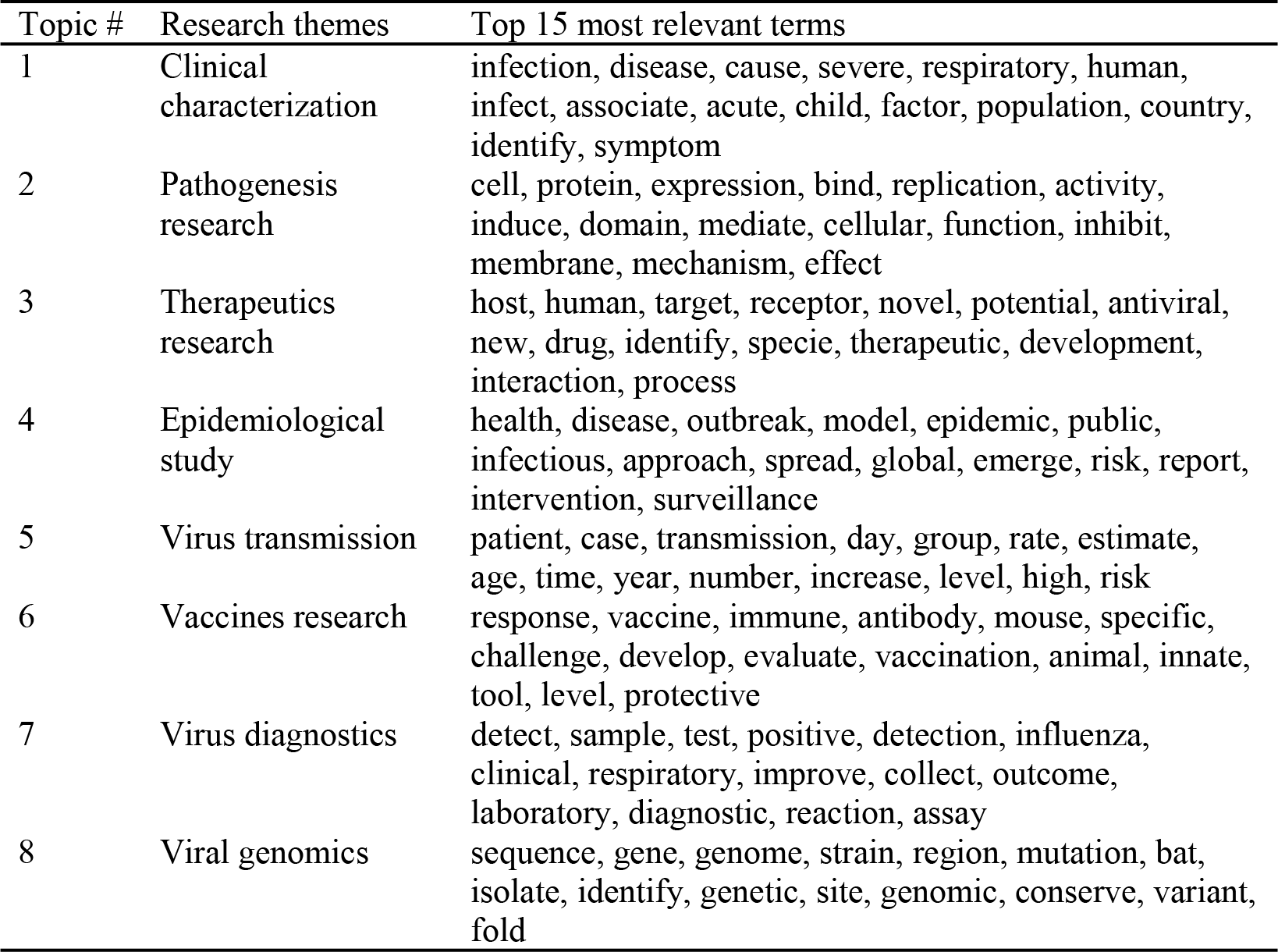
Top-15 most frequent words per topic.

| Topic # | Research themes | Top 15 most relevant terms |
| --- | --- | --- |
| 1 | Clinical characterization | infection, disease, cause, severe, respiratory, human, infect, associate, acute, child, factor, population, country, identify, symptom |
| 2 | Pathogenesis research | cell, protein, expression, bind, replication, activity, induce, domain, mediate, cellular, function, inhibit, membrane, mechanism, effect |
| 3 | Therapeutics research | host, human, target, receptor, novel, potential, antiviral, new, drug, identify, specie, therapeutic, development, interaction, process |
| 4 | Epidemiological study | health, disease, outbreak, model, epidemic, public, infectious, approach, spread, global, emerge, risk, report, intervention, surveillance |
| 5 | Virus transmission | patient, case, transmission, day, group, rate, estimate, age, time, year, number, increase, level, high, risk |
| 6 | Vaccines research | response, vaccine, immune, antibody, mouse, specific, challenge, develop, evaluate, vaccination, animal, innate, tool, level, protective |
| 7 | Virus diagnostics | detect, sample, test, positive, detection, influenza, clinical, respiratory, improve, collect, outcome, laboratory, diagnostic, reaction, assay |
| 8 | Viral genomics | sequence, gene, genome, strain, region, mutation, bat, isolate, identify, genetic, site, genomic, conserve, variant, fold |

### Semantic relationships between topics

It’s common that one article discusses more than one topic, i.e., the topics are not isolated but semantic-related. To identify the semantic relationships between topics, we assigned each article two topics with the highest probabilistic proportion, measured the relationships between two topics by using their co-occurrence statistics, and showed them vividly in a direct chord diagram. As shown in Fig. 3, the topic ‘clinical characterization’ has strong relationships with all the other seven topics, especially the virus transmission, indicating the basic role of the clinical characterization research. Besides, there is a very strong semantic relationship between any two of ‘clinical characterization’, ‘pathogenesis research’ and ‘therapeutics research’. This seems reasonable and suggests that these three topics can provide each other with a wealth of information. The topic ‘epidemiological study’ shares close relationships with ‘clinical characterization’ because clinical characterization investigation is an important part of epidemiology. Meanwhile, close relationships also exist between ‘epidemiological study’ and ‘therapeutics research’ or ‘virus transmission’, probably due to the importance of epidemiology in the therapeutics discovery and virus transmission blocking. Furthermore, it is impressive that topics ‘virus diagnostics’, ‘viral genomics’ and ‘vaccines research’ are relatively independent, interacting less with other topics.

**Fig 3.**
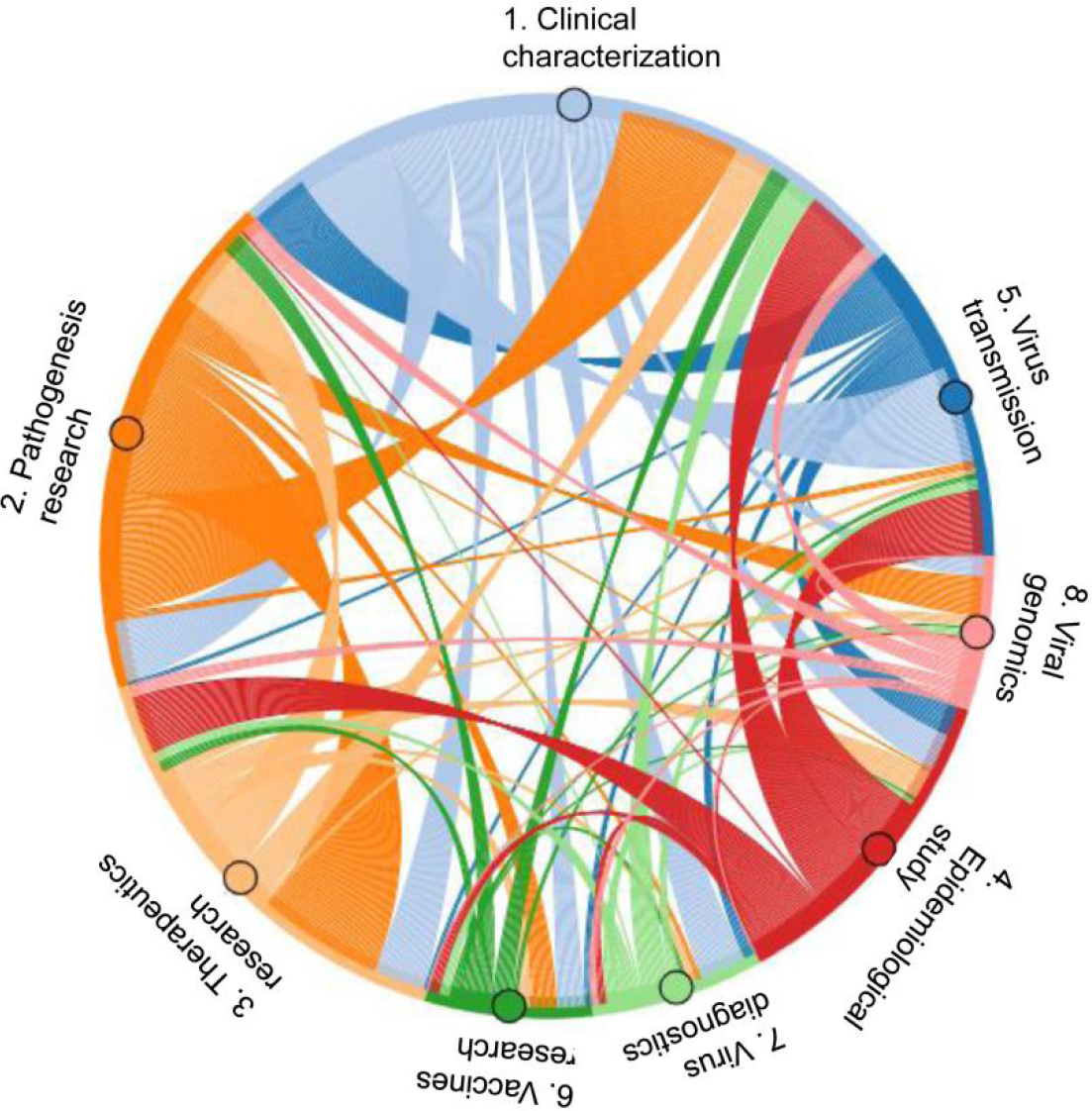
The chord diagram of semantic relationships between topics. Each band represents a topic with its width representing the number of articles with that topic being their dominant or second topic. Each ribbon represents a relationship connecting a dominant topic (bands around the periphery and ribbons arising from the bands of the same color) to the next topic (endpoint of the ribbon with different color than the adjacent band). The width of the ribbons represents the number of relationships with that same co-occurrence as the dominant and second topics.

### Comparison of research hotspots between COVID-19 and other coronavirus infections

To evaluate the current state of knowledge on COVID-19, we compared the topic distributions of 1,482 COVID-19 related articles with that of 33,610 other CoVs infections related articles. Fig. 4 shows the overall occurrence of each research topic across the COVID-19 research with most prevalent topics attributable to ‘clinical characterization’ (27.94%), ‘epidemiological study’ (27.80%), and ‘virus transmission’ (25.03%), while other CoV infections to ‘clinical characterization’ (25.97%), ‘pathogenesis research’ (18.88%), and ‘therapeutics research’ (16.51%). We noticed that topics ‘epidemiological study’ and ‘virus transmission’ show greatly larger proportions on COVID-19 than on other CoV infections. In comparison, the popularities of topics ‘pathogenesis research’, ‘therapeutics research’, ‘virus diagnostics’, ‘vaccines research’ and ‘viral genomics’ on COVID-19 are significantly lower than those on other CoV infections, accounting for less than 10% or even 4% of all the COVID-19 publications. These results indicate that research hotspots on COVID-19 are not consistent with other CoV infections, and some topics are very attractive to the research community at present.

**Fig 4.**
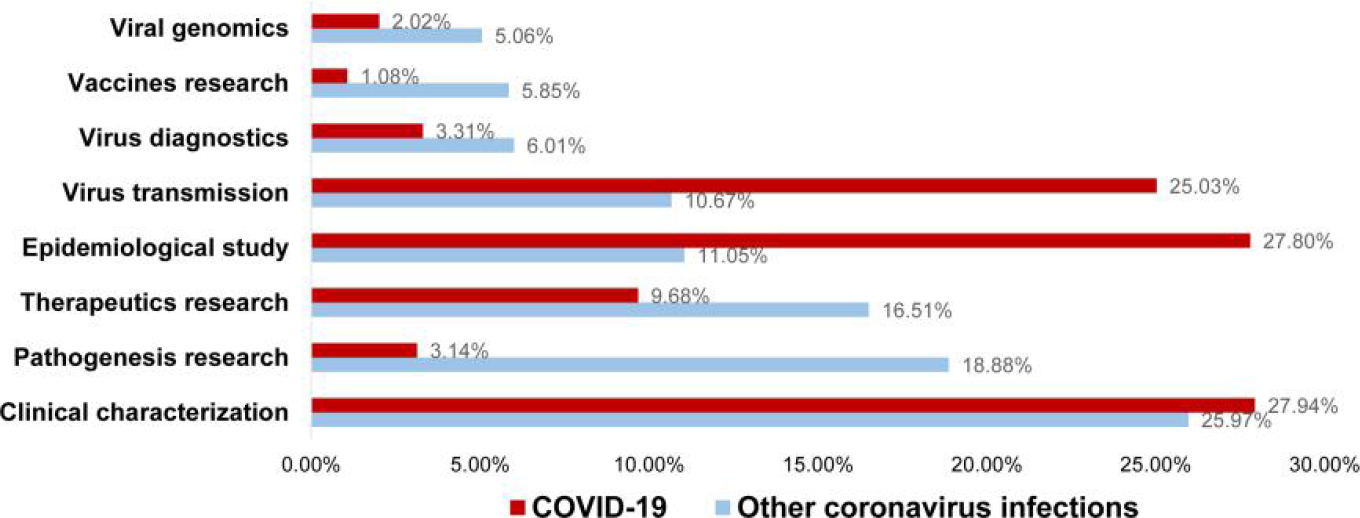
Topic distribution of COVID-19 and other coronavirus infections.

## Discussion

Here we applied LDA to the abstracts of 35,092 CoVs-related research articles up to March 20, 2020. Results of this exploration present a comprehensive overview and an intellectual structure of the coronavirus research. We identified eight major research topics, analyzed their semantic relationships, and compared the topic distributions between the evolving COVID-19 research and other CoVs research. This study found more frequent appearances of clinical characterization, epidemiological study and virus transmission in COVID-19 studies. However, there still is a lack of enough research on pathogenesis, therapeutics, virus diagnostics, vaccines and viral genomics, informing the directions of scientific development in this knowledge domain.

Research hotspots in our results do show similarities with the selected knowledge gaps in “A COORDINATED GLOBAL RESEARCH ROADMAP: 2019 NOVEL CORONAVIRUS” published by the WHO (8), which is consensus of over 400 experts from across the world. Six of our eight distinct topics (‘clinical characterization’, ‘therapeutics research’, ‘epidemiological study’, ‘virus transmission’, ‘virus diagnostics’ and ‘vaccines research’) were listed as research priorities in the roadmap. This suggests that our topic modeling successfully captured the hotspots of CoV research, thus giving us a chance to quantify how much has been investigated about COVID-19.

There exists an obvious phenomenon whereby the proportions of some topics of COVID-19 are smaller or larger than the proportions of topics based only on other COVID-19 infections. Normally, for viruses that broke out earlier, certain topics can be attractive and widely discussed. In the case of topics with high proportions, we consider that they have been elaborated and specified by research community, such as the topics ‘clinical characterization’, ‘pathogenesis research’, and ‘therapeutics research’ in other CoVs research. While as for the new virus SARS-CoV-2, the research topic distribution is in a state of dynamic change. Therefore, comparison between the ‘old’ and ‘new’ viruses could clearly highlight the knowledge gaps between them.

This work suggests that research needs to be focused on pathogenesis, therapeutics, virus diagnostics, vaccines and viral genomics for COVID-19, which could then necessitate national-level support for ongoing research, if required. Pathogenesis research is usually important but time-consuming. We believe such studies will be available in the future. The more important and urgent topic is therapeutics research, which accounts for only 9.68% in COVID-19 research, much lower than 18.88% in other CoV infections research. With the number of infection cases increasing rapidly, therapeutics research will be more and more urgently needed. In addition, virus diagnostics, vaccines and viral genomics show small proportions in all CoV infections research, but even smaller in the field of COVID-19. This shows how little we know about CoVs. It is time to translate research findings into effective measures such as a diagnostics, vaccine or effective therapeutic options, as with other priority diseases (22).

Similar to the study of Md Mahbub Hossain MBBS (10), we did not identify any topic about social, economic, political, psychological, or cultural consequences of COVID-19 as well as other CoV infections. The WHO roadmap has noticed this and taken ‘social sciences in the outbreak response’ as a priority. The social science research community should act to assess the impact of a major public health crisis like COVID-19 on the complex network of modern societies (23, 24).

The topic modeling analysis identified meaningful semantic relationships between research topics, probably because of shared information or shared methods. The clinical characterization research is almost equally important for every research topic area as a link among clinical, macroscopic and microscopic research. The epidemiological study plays a key role in the blockage of virus transmission. The study of viral genomics is relatively isolated, but it is important to identify pathogens and their variants at an early stage of virus outbreak. In response to the SARS-CoV-2 outbreak, scientists in China immediately started to research the source of the new coronavirus, and rapidly published the first genome of SARS-CoV-2 on 10 January 2020, which greatly promoted virus diagnostics and epidemiological research (25). By highlighting such theme interactions within coronavirus research, we hope to inspire new collaborations among researchers, by sharing data, information, technologies and ideas. The fight against the COVID-19 pandemic will benefit from active efforts to cross fertilize.

This present work has several limitations that must be acknowledged to apprehend the findings. First, as a computer model, topic modeling has difficulties with understanding nuances and subtext. We identified eight major topics with frequent appearances, whereas some specific topics such as virus natural history and human-animal interface were buried due to low proportions. Future work will attempt to develop an approach to detect and visualize the conceptual sub-domains. Second, it is also necessary to be aware that the granularity of terms used to label topics may vary a little for different topics. We have tried our best to keep them at the same level through carefully curating the top words in each topic, as well as reviewing text intention of the corresponding publications. Third, the LDA model only provides a cross-sectional profile of the coronavirus research, which is methodologically different from academic disciplines such as evidence-based medicine and clinical assessment that synthesizing evidence centered on a specific question. But this study may facilitate these disciplines by informing what is already available and what is urgently required for the COVID-19 research. Future research could also study the occurrence of topics over time and analyze links to historic events and virus characteristics, in order to better understand possible temporal patterns.

Notwithstanding its limitations, this work is the first to thoroughly assess research output of coronavirus research in statistical perspective. The present study provided machine-extracted research topics in various contexts based on massive number of articles over a long period. Such a bottom-up approach can complement the existing manual content analysis techniques. It can also help to find the latent semantic relationships between topics and enrich the research landscape.

In conclusion, this study specifically shows a holistic picture of the current research in response to the outbreak of COVID-19. We conducted a topic-based bibliometric study to address two questions “What is the CoVs research interested in?” and “Where are we with our research about COVID-19?”. An LDA-based model was used to identify the hotspots and their difference between COVID-19 and other CoV infections in a quantitative way. Generally, eight main research areas are identified, i.e., clinical characterization, pathogenesis research, therapeutics research, epidemiological study, virus transmission, vaccines research, virus diagnostics, and viral genomics. The results also indicate that studies in diagnostics, therapeutics, vaccines, viral genomics, and pathogenesis are urgently needed to minimize the impact of the COVID-19 outbreak. As COVID-19 can be considered a recent emerged disease, we characterize a ‘snapshot’ of this field at an early stage in its development. Our findings can potentially aid governments and the research community in understanding recent development of the research field, optimizing research topic decision, and monitoring new scientific or technological activities.

## Data Availability

All the data are publicly available (https://www.kaggle.com/allen-institute-for-ai/CORD-19-research-challenge/).

## Acknowledgements

This work was supported by the National Key R&D Plan of China (grant number 2018YFC1315405 to XC), the National Natural Science Foundation of China (grant number 71704031 to LL), and the fund from Guangzhou Institute of Pediatrics/Guangzhou Women and Children’s Medical Center (grant numbers IP-2019-017 to GL, KCP-2016-002 to HL).

